# Pitfalls in EEG analysis in patients with non-convulsive status epilepticus

**DOI:** 10.1101/2020.10.02.20205583

**Authors:** Ying Wang, Ivan C Zibrandtsen, Richard HC Lazeron, Johannes P van Dijk, Xi Long, Ronald M Aarts, Lei Wang, Johan BAM Arends

## Abstract

**Objective:** Electroencephalography (EEG) interpretations through visual (by human raters) and automated (by computer technology) analysis are still not reliable for the diagnosis of non-convulsive status epilepticus (NCSE). This study aimed to identify typical pitfalls in the EEG analysis and make suggestions as to how those pitfalls might be avoided.

**Methods:** We analyzed the EEG recordings of individuals who had clinically confirmed or suspected NCSE. Epileptiform EEG activity during seizures (ictal discharges) were visually analyzed by two independent raters. We investigated whether unreliable EEG visual interpretations quantified by low inter-rater agreement can be predicted by the characteristics of ictal discharges and individuals’ clinical data. In addition, the EEG recordings were automatically analyzed by in-house algorithms. To further explore the causes of unreliable EEG interpretations, two epileptologists analyzed EEG patterns most likely misinterpreted as ictal discharges based on the differences between the EEG interpretations through the visual and automated analysis.

**Results:** Short ictal discharges with a gradual onset (developing over 3 seconds in length) were liable to be misinterpreted. An extra 2 minutes of ictal discharges contributed to an increase in the kappa statistics of > 0.1. Other problems were the misinterpretation of abnormal background activity (slow wave activities, other abnormal brain activity, and the ictal-like movement artifacts), continuous interictal discharges, and continuous short ictal discharges.

**Conclusion:** A longer duration criterion for NCSE-EEGs than 10 seconds that commonly used in NCSE working criteria is needed. Using knowledge of historical EEGs, individualized algorithms, and context-dependent alarm thresholds may also avoid the pitfalls.

## 1. Introduction

Non-convulsive status epilepticus (NCSE) is characterized by its inconspicuous motor signs with prolonged electrographic seizure activities ^1^. Given the subtle and variable clinical presentations, electroencephalography (EEG) that confirms ictal discharges (epileptiform EEG activity during a seizure) is an essential diagnostic tool of NCSE. The reliability of the diagnostic tool can be assessed by the interrater agreement of visual EEG interpretation ^2^. A low agreement could indicate the presence of certain EEG misinterpretations and therefore less reliable diagnosis of NCSE ^3^.

Currently, NCSE-EEG interpretations through visual analysis by human raters and automated analysis by computer technology are still not very reliable ^4–10^. The Salzburg Consensus Criteria (SCC) ^11,12^ achieved a reasonably high accuracy and inter-rater agreement among human raters for the diagnosis of NCSE ^13,14^. However, the accuracy and agreement are relatively low when human raters cannot assess the effect of intravenous antiepileptic drugs on the EEG of individuals during NCSE and when raters interpret short EEG recordings ^5,15,16^. The visual EEG interpretation relies heavily on subjective judgments based on human raters’ experience and knowledge about the characteristics of ictal discharges such as location, morphology, frequency, and persistence. EEG-readers who are inexperienced for NCSE-EEG patterns are more liable to misinterpret EEG, which results in an unreliable NCSE diagnosis. A previous study ^10^ summarized several pitfalls in the EEG interpretations of intensive care unit (ICU) patients with NCSE, including misinterpreting artifacts as ictal discharges, assuming that the stereotypical patterns of ictal discharges are observed on the same subject, and assuming that a dichotomy exists between ictal and interictal discharges (epileptiform EEG activity between seizures) in patients with encephalopathy. On the other hand, automated EEG analysis can assist clinicians and care-givers in the detection of ictal discharges because of its greater time efficiency and more objective judgments than visual analysis ^6–9^. However, pitfalls of EEG misinterpretations (misclassifications) also exist in automated analysis ^8,9^: the misclassification of pre-ictal discharges (epileptiform EEG activity before a seizure), post-ictal discharges (epileptiform EEG activity after a seizure), high-frequency artifacts, or similar background EEG activity. These misinterpretations of EEG by both human raters and computer technology could lead to wrong diagnoses of NCSE and cause serious consequences ^17,18^.

EEG-readers need to be alert to potential pitfalls in EEG analysis for reliable NCSE diagnosis. Nevertheless, studies on the pitfalls in the visual and automated EEG analysis for NCSE are relatively scarce ^4,10^. This research therefore addresses the pitfalls of unreliable EEG interpretations in the visual and automated EEG analysis for NCSE patients with chronic epilepsy and brain development disorders. We explored whether the reliability of EEG interpretation can be predicted by the characteristics of ictal discharges and the clinical data of patients. Moreover, we further identified potential pitfalls which could cause EEG misinterpretations and therefore low reliability in the NCSE diagnosis. Strategies to avoid the pitfalls were also proposed in this study.

## 2. Materials and methods

### 2.1. Study design

This is a retrospective explorative study approved by the Medical Research Ethics Committee of Kempenhaeghe in the Netherlands. We retrieved and analyzed the EEG and the clinical records of 30 subjects with preceding seizures between 2008 and 2016 in this study. Twenty of the 30 subjects’ data were recorded when they had clinically confirmed NCSE, which was diagnosed based on the clinical information, such as clinical signs, response to treatment, and EEG recordings. The other ten subjects’ data were recorded during clinically suspected NCSE, that is, the subjects were first suspected to have NCSE based on clinical signs, but the suspicion was disconfirmed based on the response to treatment and EEG recordings. We included subjects < 65 years and excluded EEG recordings with excessive artifacts and those of subjects from whom we did not receive permission (from themselves and/or their legal guardians) to use their recordings for scientific research.

The study had three phases. (1) Subjects’ data were screened according to the inclusion and exclusion criteria, and the clinical background was evaluated by an experienced epileptologist. (2) Ictal discharges in the EEG recordings were annotated by two independent raters. (3) Pitfalls in the visual and automated EEG analysis were investigated.

### 2.2. Data sources

The EEG recordings were acquired by three different systems (BrainRT equipment from Onafhankelijke Software Groep, EEG Stellate from Natus Europe GmbH, and Micromed from SIGMA Medizin-Technik GmbH) at Kempenhaeghe. The sampling rate of the EEG recordings was 100 Hz, 200 Hz, or 256Hz. EEG electrodes were positioned in the 10-20 system. For the algorithm development of the automated EEG analysis, 21 electrodes were used (19 electrodes in the 10-20 system, plus electrodes F9 and F10). All recordings were continuous except in nine subjects, where 5 minutes per hour were stored when no clinical abnormalities were recorded. From each subject’s clinical data and the corresponding clinical report of EEG recordings, we collected information about age, sex, intellectual disability level, sleep status, preexisting epileptic encephalopathy (defined as severe brain dysfunction at early age), clinical signs during NCSE, and seizure history.

### 2.3. EEG visual analysis and inter-rater agreement

The raters were asked to focus on the EEG visual analysis without checking corresponding videos, as NCSE is known by its difficult discrimination from normal behavior, and the fact that different patients show variable clinical presentations. Annotations of ictal discharges were made with the assist of the open-source software “EDFbrowser” ^19^. Four types of EEG montages were available during the visual analysis, including longitudinal bipolar montage, transverse bipolar montage, average montage, and original archived data montage.

The raters annotated ictal discharges and labeled their characteristics: duration, certainty, onset location, onset visibility, and morphology patterns according to the pre-defined criteria shown in *table 1*. As NCSE takes a relatively long time to develop and disappear, we set a minimum of 20 seconds as the duration criterion for the EEG pattern. The presence of ictal discharges was primarily assessed according to the combination of both SCC and the American Clinical Neurophysiology Society’s Standardized Critical Care EEG Terminology (ACNS) criteria ^11,12,20^. The raters categorized the certainty of their annotations as either definite or possible ictal discharges, and the rest of the recordings was categorized as episodes without ictal discharges. In addition, the ictal discharges were labeled either generalized or focal according to their onset location, and their onset visibility were labeled either sudden or gradual according to the ACNS criteria. Five categories were used to describe the dominant morphology patterns: three of five categories—“Spike Wave”, “Wave”, and “Fast Spike” pattern—were summarized from several terms in ACNS criteria, and the other two empirical categories were added—“Seizure-related EMG Artifacts” ^21^, and “Unknown Type”.

**Table 1.** Annotation criteria of ictal discharges.

|  |  | Definite ictal discharges | Possible ictal discharges | Not ictal discharges |
| --- | --- | --- | --- | --- |
| Duration | | $\geq 20$ seconds | | $< 20$ seconds |
| Certainty |  | - SW discharges $> 2.5$ Hz<br>- Other-pattern evolving discharges $> 4$ Hz | - SW discharges $\leq 2.5$ Hz<br>- Other-pattern evolving discharges $\leq 4$ Hz<br>- RDA $> 0.5$Hz | Rest EEG activity |
| Onset location | Generalized | Occupying $>$ half of the longitudinal bipolar channels | | |
| | Focal | Occupying ≤ half of the longitudinal bipolar channels | | |
| Onset visibility | Sudden | Developing from absent $\leq 3$ seconds | | |
| | Gradual | Developing from absent $> 3$ seconds | | |
| Morphology patterns | Spike Wave | SW discharges or RDA with superimposed repetitive sharp waves or spikes (RDA+S) |  |  |
|  | Wave | RDA |  |  |
|  | Fast Spike | Spiky or sharp periodic discharges |  |  |
|  | Seizure-related EMG Artifacts | Electromyography artifacts caused by seizures |  |  |
|  | Unknown Type | Other patterns than the above four |  |  |

EEG: electroencephalography; EMG: electromyography; RDA: rhythmic delta activity; SW: spike-and-wave or sharp-and-wave.

We summarized the annotated ictal discharges by the two raters via an annotation code (*appendix A*), and estimated the inter-rater agreement. Cohen’s kappa was calculated to assess the agreement on episodes with and without ictal discharges. Fleiss’ kappa was estimated to assess the agreement on episodes with definite ictal discharges, with possible ictal discharges, and without ictal discharges. The 95% confidence interval (CI) of the kappa statistics were also calculated. The level of the agreement was interpreted according to the suggestions of Landis and Koch ^22^.

### 2.4. Investigation of unreliable EEG interpretations through visual analysis

We applied linear regression models to determine whether the reliability of EEG interpretations, quantified by the inter-rater agreement, was influenced by the characteristics of ictal discharges and the clinical data of subjects. Two models with Cohen’s kappa and Fleiss’ kappa as the dependent variables were fitted. The independent variables were the characteristics of ictal discharges (duration, certainty, onset location, onset visibility, and morphology patterns) and the clinical data (age, sex, intellectual disability level, sleep status, preexisting epileptic encephalopathy, clinical signs during NCSE, and seizure history). The analysis was implemented in SPSS Statistics version 25. A two-tailed p-value < 0.05 indicated a significant linear relationship.

### 2.5. Analysis of EEG misinterpretations through visual and automated analysis

Comparing the EEG interpretations from visual and automated analysis can help us point out the hidden pitfalls. Therefore, we developed a customized multimodal viewer (a Matlab graphical user interface shown in *figure 1*) that presents the EEG recordings, the annotated ictal discharges by raters, and the EEG signal classification results generated by our in-house automated analysis system. The automated analysis system included pre-processing, feature extraction, and synthetic three-class classification (more details can be found in *appendix B*). The system mainly analyzed features in time-frequency domain and built a synthetic three-class classifier to classify EEG epochs into three categories: ictal discharges, suspicious activity, and normal activity. The suspicious activity indicated the misinterpreted EEG signals (signals misclassified as ictal discharges) in the automated analysis. In the lower panel of the multimodal viewer (*figure 1*), the distribution of the three categories were presented in units of 10 seconds.

**Figure 1.**
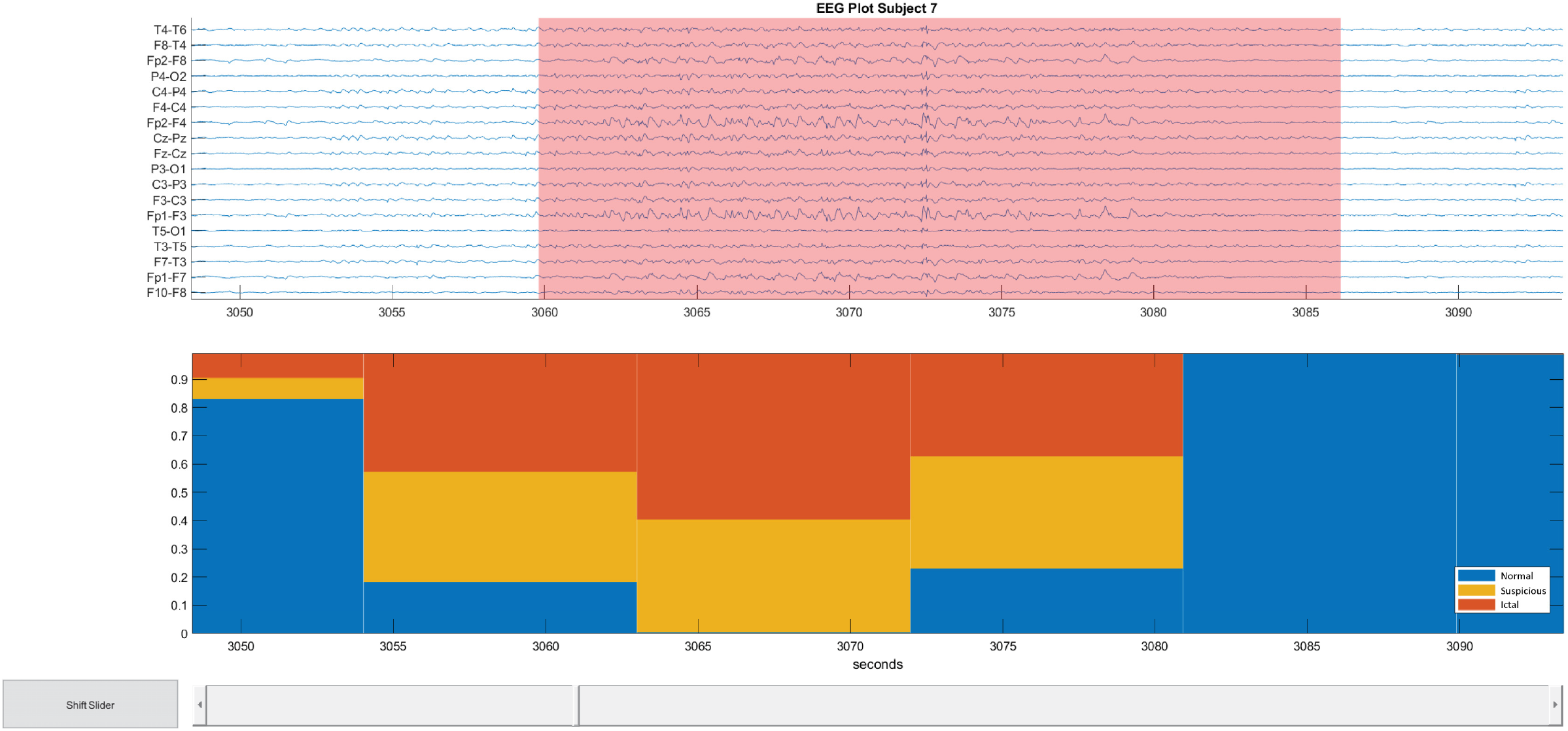
Viewer for EEG recording and classification results. The upper panel in the viewer presents an EEG recording in the longitudinal bipolar montage. Pink backgrounds indicate ictal discharges. The lower panel shows the stacked percentages of the classification results. Each bin in the bar plot indicates 10 seconds. The upper and lower panels are linked, and both of them are able to be zoomed in or out. EEG: electroencephalography.

Two epileptologists investigated and discussed the pitfalls of EEG misinterpretations for each subject. They used the multimodal viewer to compare the EEG interpretations through the visual and automated analysis. Meanwhile, they checked the corresponding clinical data and inter-rater agreement. The pitfalls of EEG misinterpretations in the automated analysis were investigated among the EEG episodes without annotated ictal discharges but automatically classified as definite or suspected ictal discharges. The pitfalls of EEG misinterpretations in the visual analysis were investigated among the EEG episodes with annotated ictal discharges but automatically classified as normal activity

## 3. Results

### 3.1. Subjects

The subject flow through the three study phases is shown in a recruitment tree (*figure 2*). In the first phase, two subjects were excluded, and two were moved from the diagnosed NCSE group to the suspected NCSE group. In the second phase, the EEG recordings of two subjects in the NCSE group and nine subjects in the suspected NCSE group did not include any agreed annotated ictal discharges. All 16 subjects in the NCSE group were included in the third phase for the statistical analysis. Fourteen of the 16 subjects were used in the automated EEG analysis system development. In addition, two, nine, and five of the 16 subjects’ EEG recordings in the NCSE group were sampled at 100 Hz, 200 Hz, and 256 Hz, respectively. In the suspected NCSE group, one subject’s recording was sampled at 200 Hz, and the others were at 256 Hz.

**Figure 2.**
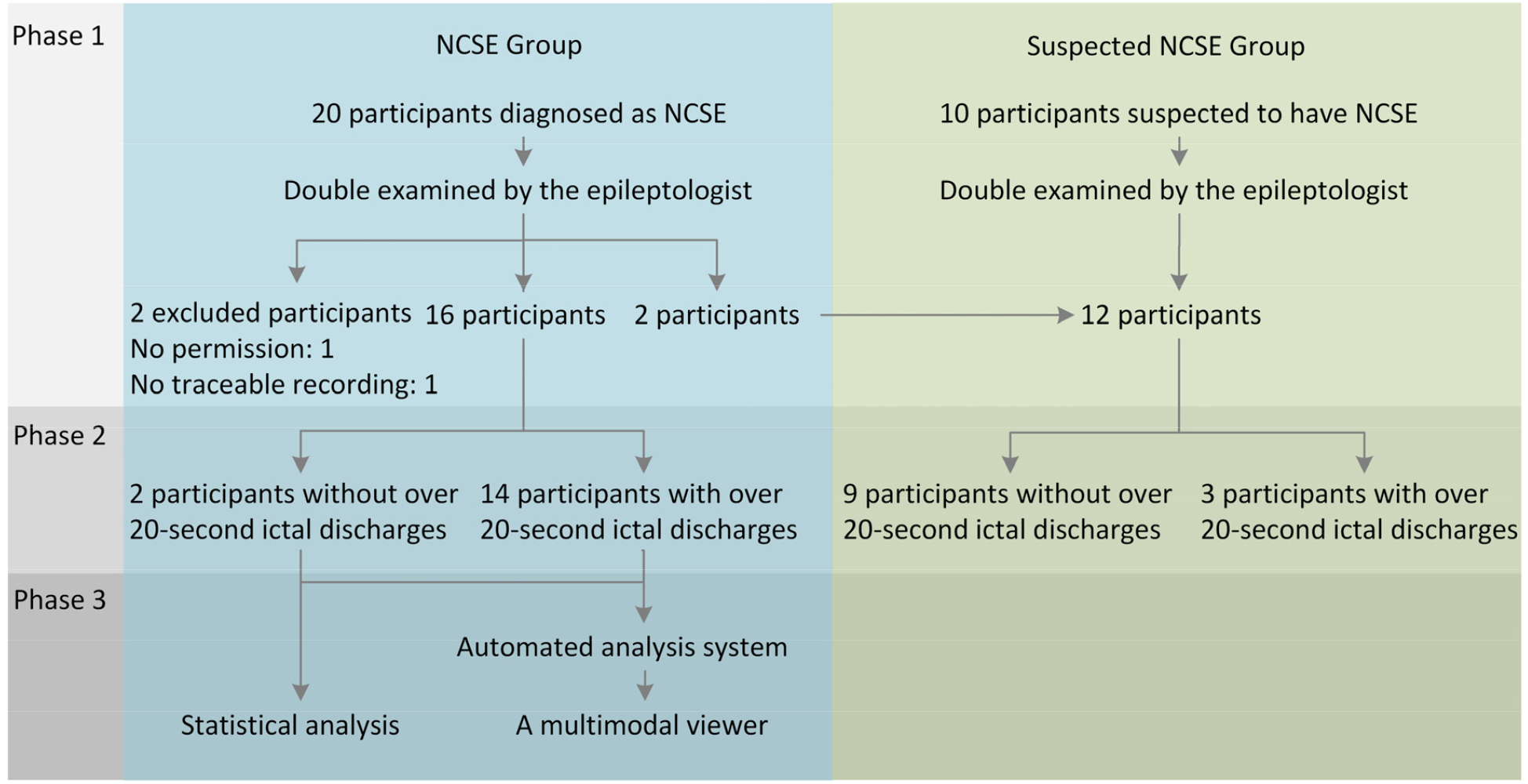
Recruitment tree.

In the first phase, the median age of the 16 NCSE subjects was 21 years, ranging from 6 to 43 years, and the median age of the 12 suspected NCSE subjects was 19 years, ranging from 4 to 61 years. *Figure 3* presents an overview of the demographics and the clinical characteristics of the subjects. Of note, two of 16 NCSE subjects had preexisting epileptic encephalopathy, whereas in the suspected NCSE group, the subjects with and without preexisting epileptic encephalopathy were equally distributed. Moreover, the EEG recording duration in the suspected NCSE group is generally shorter than that in the NCSE group. A more comprehensive list of the clinical signs, seizure histories, and other characteristics of individual NCSE and suspected NCSE subjects are provided in *appendix C*.

**Figure 3.**
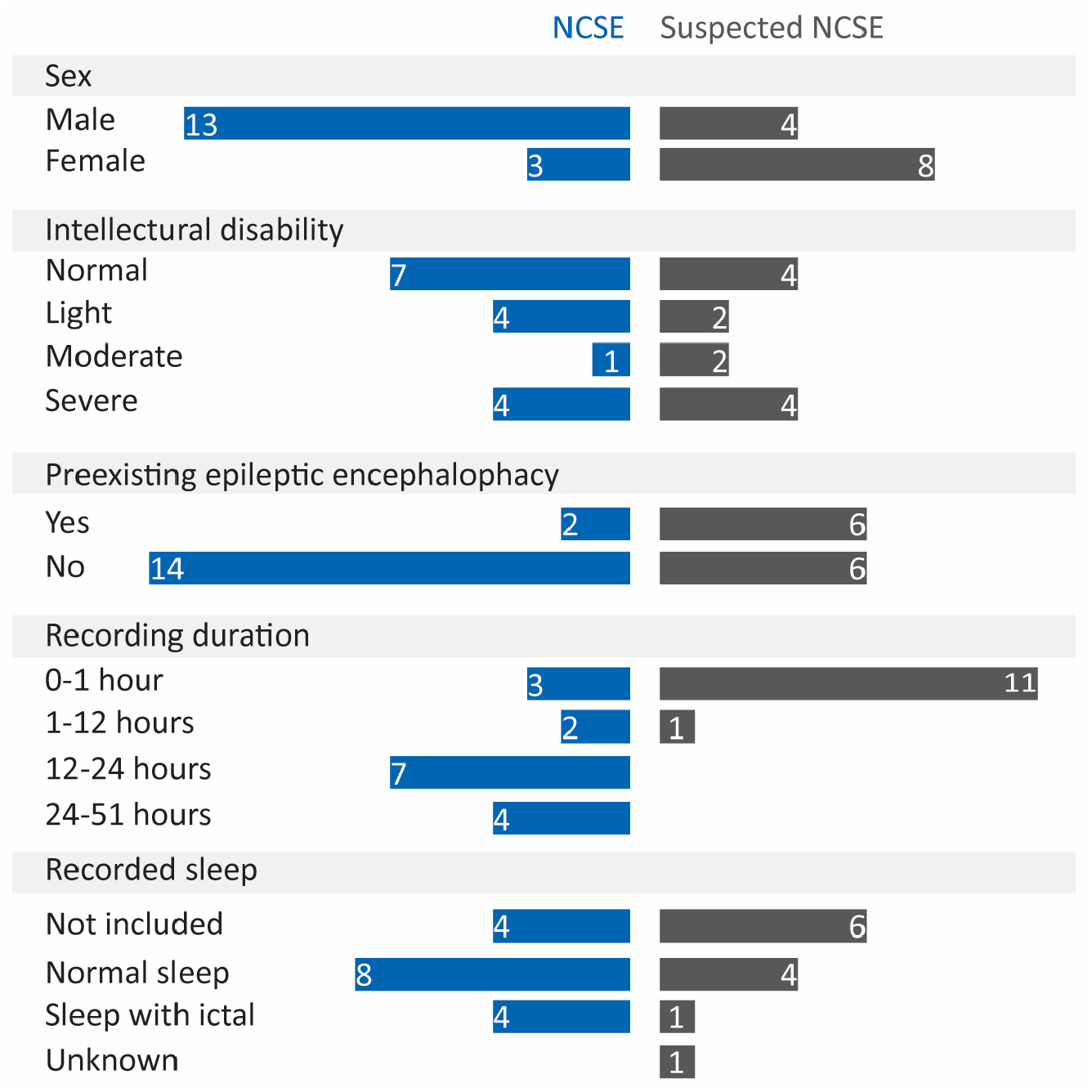
The overview of the demographics and the clinical characteristics of the subjects in the NCSE and suspected NCSE groups. NCSE: non-convulsive status epilepticus.

### 3.2. Inter-rater agreement of visual EEG interpretations

In the NCSE group, the inter-rater agreement for both two categories (episodes with ictal discharges and without ictal discharges) and three categories (episodes with possible ictal discharges, with definite ictal discharges, and without ictal discharges) was moderate, with Cohen’s kappa = 0.53 and 95% CI [0.37, 0.69], and Fleiss’ kappa = 0.41 and 95% CI [0.25, 0.57], respectively. The inter-rater agreement of the individual subjects are shown in *appendix D*. In the suspected NCSE group, the inter-rater agreement was poor (Cohen’s kappa = 0; Fleiss’ kappa = −0.08); hence, we did not further analyze the EEG recordings in this group.

### 3.3. Annotated ictal discharges in the NCSE group

In the NCSE group, 338 ictal discharges were annotated among approximately 183 hours of EEG recordings. The annotated ictal discharges lasted approximately 14.7 hours in total. The ictal discharges were summarized (*figure 4*) according to their characteristics: the certainty, onset location, onset visibility, and morphology patterns. The definite and possible ictal discharges each accounted for similar proportions. With respect to the onset location, generalized ictal discharges reached major proportions. In addition, the ictal discharges with a sudden onset, developing from absent in less than or equal to 3 seconds, accounted for a slightly higher proportion than the ictal discharges with a gradual onset, developing from absent in more than 3 seconds. Morphologically, the vast majority (66%) of the ictal discharges had a “Spike Wave” pattern.

**Figure 4.**
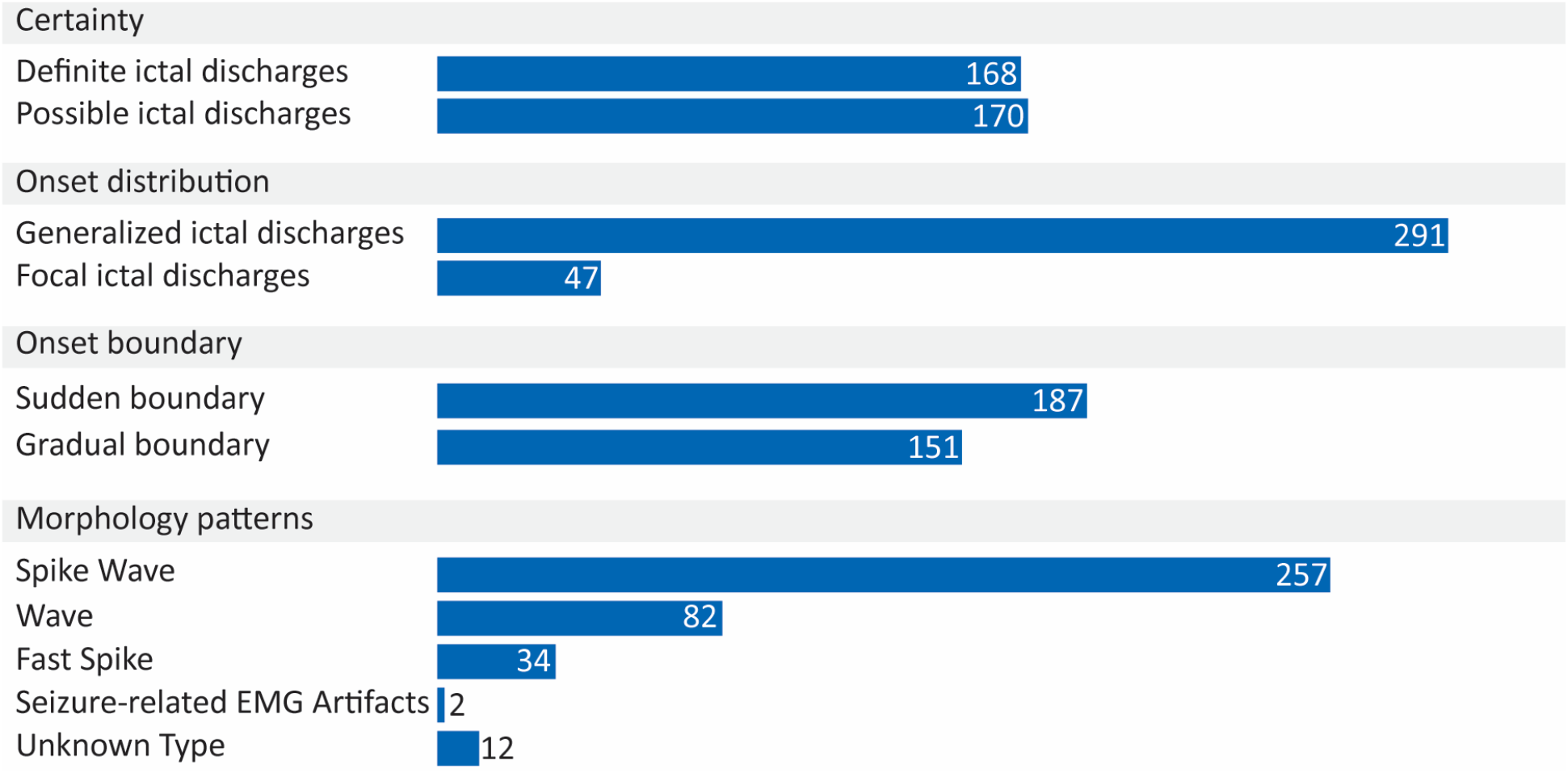
The number of the ictal discharges categorized by their characteristics.

### 3.4. Pitfalls of unreliable EEG interpretations through visual EEG analysis

Linear models showed a significant relationship between the average duration of the ictal discharges with a gradual onset and inter-rater agreement measured by the kappa statistics. For Cohen’s kappa, β=0.001, F(1, 14) = 10.861, p = 0.005, and the independent variable (the average duration) explained 43.7% of the variability. For Fleiss’s kappa, β=0.001, F(1, 14) = 12.89, p = 0.003, and the independent variable explained 44.2% of the variability. In other words, an extra 2 minutes of the ictal discharges with a gradual onset contributed to an increase in the kappa statistics of > 0.1. This implies that human raters interpreted ictal discharges with a gradual onset less reliably when the duration was shorter. Using a short duration criterion in annotating these ictal discharges could be a pitfall in visual EEG analysis (*table 2*).

**Table 2.** Pitfalls in EEG interpretations.

|  |
| --- |
| **Pitfalls of unreliable EEG interpretations through visual analysis** |
| Using a short duration criterion in annotating ictal with a gradual onset |
| **Pitfalls of EEG over-interpretations** |
| Misinterpretation of abnormal background activity: |
| - Misinterpretation of slow wave activities observed in a dysfunctional brain or in a drowsy stage;<br>- Assuming that the EEG background of individual subjects was consistently normal, and over-interpreting the abnormalities as ictal;<br>- Misinterpretation of the envelope of ictal-like movement artifacts leads to false alarms in computer-assisted EEG analysis |
| Misinterpretation of continuous interictal discharges |
| Misinterpretation of continuous short ictal discharges |

EEG: electroencephalography.

### 3.5. Expert opinion of the reasons of misinterpretations by visual and automated EEG analysis

The epileptologists summarized the pitfalls of EEG misinterpretations using the multimodal viewer. The pitfalls were the misinterpretation of the following:

- abnormal background activity (*figure 5A-C*),
- continuous interictal discharges (*figure 5D*),
- continuous short ictal discharges (*figure 5E*) whose duration were shorter than 20 seconds.

**Figure 5.**
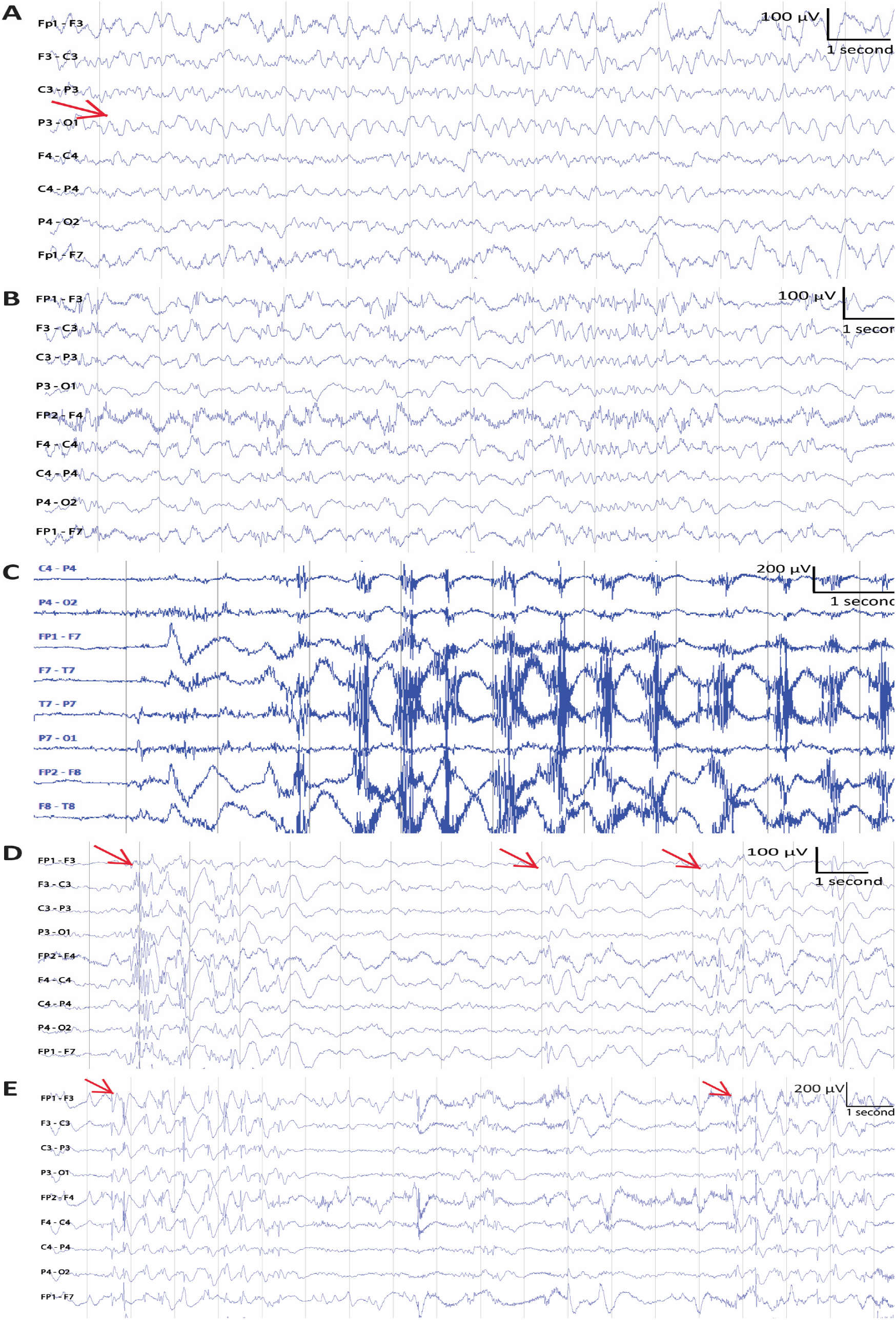
Abnormal background activity. (A) Slow wave activities marked by an arrow. (B) Other abnormal brain activity. (C) Ictal-like movement artifacts. (D) Several interictal discharges. The onsets of the interictal discharges are marked by arrows. (E) Short ictal discharges. The onsets of two short ictal discharges are labeled by arrows.

The abnormal background activity was categorized in three types: slow wave activities (*figure 5A*) regularly observed in a dysfunctional brain or in a drowsy stage, other abnormal brain activity (*figure 5B*) whose appearance was subject-dependent, and ictal-discharge-like movement artifacts caused by rhythmic movements, such as repetitive chewing movements (*figure 5C*). The pitfalls of misinterpretations are summarized in *table 2*, and the remarks by the epileptologists are provided in *appendix E*.

## 4. Discussion

A reliable and correct EEG interpretation to diagnose NCSE is currently still difficult from visual as well as automated analysis ^5,18,23^. In this retrospective study, we visually and automatically analyzed NCSE-EEG recordings, and the inter-rater agreement in the NCSE group was moderate (Cohen’s kappa = 0.53 and Fleiss’ kappa = 0.41), consistent with the findings by Goselink et al. ^5^. We found that using a short duration criterion for ictal discharges with a gradual onset contributed to an unreliable EEG interpretation. Moreover, we pointed out other facts for EEG misinterpretation: abnormal background activity, continuous interictal discharges, and continuous short ictal discharges, which extended the findings of a previous study ^10^.

One pitfall was to use a short ictal duration criterion for ictal discharges with a gradual onset. In this study, we set 20 seconds as the duration criterion for ictal discharges, but even then the inter-rater agreement was still low. The 10-second criterion in SCC ^12^, defined according to commonly used EEG reading duration via software, could be too short to support the reliability of visual EEG interpretations in clinical practice. To increase the reliability of NCSE diagnosis and conform to the natural course of the disease, a longer duration criterion can be recommended in future, especially when EEG-readers rate ictal discharges with a gradual onset. However, using a too long duration criterion might increase the risk of ignoring intervals carrying essential information. Further research should be undertaken into optimal length. Until now, automated EEG analysis algorithms for NCSE have regularly used short (e.g., 3 seconds) EEG signals as a classification epoch, but the epoch duration is too short to be used in reliably detecting the ictal discharges for NCSE diagnosis according to the earlier discussion. To be consistent with the way clinicians diagnose NCSE, future studies about automated EEG analysis algorithms should focus on ictal discharge detection using longer-duration EEG epochs.

The pitfalls of EEG misinterpretations in both visual and automated analysis were misinterpreting the following as ictal discharges: (1) abnormal background activity [(a) slow wave activities, (b) other abnormal brain activity, and (c) the ictal-like movement artifacts], (2) continuous interictal discharges, and (3) continuous short ictal discharges.

(1) For the three types of activities of the abnormal background activity:

(a) Slow wave activities are frequently observed in a damaged brain (e.g., in epileptic encephalopathies) or during drowsiness. In visual analysis, EEG readers with insufficient training in reading EEGs from a dysfunctional brain or sleep-EEG may misinterpret the EEG ^18^. In automated analysis, slow wave activities and ictal discharges could be confused because their frequency bands are similar. An advanced signal processing technique, such as extracting features in the morphological besides the time-frequency domain features ^24^, may be helpful in their distinction in future studies.

(b) The other abnormal brain activity is individually variable, and can be avoided by the use of historical EEG data. Its misinterpretation (false alarms) in automated analysis was also mentioned in the previous studies ^8,9^.

(c) The ictal-like movement artifacts could be misinterpreted in automated analysis when their repetitive movements have a similar frequency band as ictal discharges. The low-pass filter of the analysis algorithms helps filter out high-frequency components caused by muscle activities, but keeps the envelope of signals caused by the repetitive movements.

In summary, to avoid misinterpretation of abnormal background activity, we suggest reading more EEG recordings from the same subject to help human raters better recognize subject-specific ictal discharges and individualizing automated analysis algorithms.

(2) Interictal discharges are not always clearly distinct from ictal discharges, and misinterpretation of continuous interictal discharges occurs especially in patients with encephalopathy ^10^. The duration and presence of repetitive spiking or bursting activity cannot be the only criteria to identify interictal discharges by EEG readers ^25^. Further investigation of criteria for interictal discharges is needed.

(3) High concentrations of short ictal discharges can be observed in the EEG recording of a subject presenting many short and unstable epileptic activities. Raters with hyper-sensitivity may misinterpret them as ictal discharges. We would suggest that EEG readers carefully interpret such EEG recordings and consult other readers if in doubt. In addition, we recommend tuning alarm thresholds according to subject-dependent clinical practice in the automated EEG analysis for these particular recordings.

One unanticipated finding is that the inter-rater agreement in the suspected NCSE group was poor. Based on our previous discussions and the clinical data of the suspected NCSE subjects, we could assume that two factors may explain the unreliable EEG interpretations in this group. (1) Almost all the recordings were less than 1 hour, and the raters may not have had enough EEG recordings from the same subject to correctly distinguish ictal discharges from the abnormal background activity. (2) Half of the subjects had preexisting epileptic encephalopathy. The slow wave activities and the interictal discharges in the subjects with encephalopathy were probably misinterpreted as ictal discharges. Additional studies are needed to confirm the causes of the poor inter-rater agreement in the EEG interpretations of patients with suspected NCSE.

This study has several limitations. Given that the number of recordings from patients with NCSE at Kempenhaeghe is relatively small, we also included several discontinuous archived EEG recordings. These discontinuous recordings may hinder human readers and automatic algorithms from correctly interpreting EEG. Continuous recordings from more data sources should be used in future. The EEG-recordings were acquired by three systems, and their sampling rate were different [the lowest sampling rate was 100 Hz (n=2)]. The heterogeneity of the sampling rate was not expected to affect our results, such as the problem of the aliasing, because the frequency band of interest, such as the frequency range occupied by Spike Wave and Wave patterns, is much lower than 50 Hz. Nevertheless, future works should be undertaken to further confirm the influence of the different system hardware and setting-up on our results. Given that this is an explorative study, the number of recruited subjects (n = 30), EEG readers (n = 2), and research centers (n = 1) was small; hence, the generalization of the conclusions is limited. In addition, given that our study is explorative and a large population of patient requires a large sample size, the subjects included in this study were outpatients with chronic seizures and brain development disorders, and their age ranged between 4 and 61 years; thus, the conclusions do not extend beyond this population. Future work should include more subjects, especially those of neonatal age, elderly patients, patients without preexisting epilepsy, and ICU patients, and more EEG readers from different research centers for the generalizability. In the development of the automated EEG analysis algorithm, we primarily used time-frequency domain features, which limited our conclusion of the pitfalls in the automated analysis. In future, morphological features should be added to the automated analysis.

## 5. Conclusion

We visually and automatically analyzed NCSE-EEG recordings to explore the causes of EEG misinterpretations. To avoid the pitfalls in NCSE-EEG analysis, a longer duration criterion than the one suggested by the Salzburg criteria is needed. Using knowledge of historical EEGs, individualized algorithms, and context-dependent alarm thresholds may also avoid mistakes.

## Data Availability

The data that support the findings of this study are available from Academic Center for Epileptology Kempenhaeghe, Heeze, the Netherlands. Restrictions apply to the availability of these data, which were used under license for this study. Data are available from the corresponding author (Y. Wang) with the permission of Academic Center for Epileptology Kempenhaeghe, Heeze, the Netherlands.

## Acknowledgements

This work is part of the research program BrainWave with project number 14714, which is (partly) financed by the Netherlands Organization for Scientific Research (NWO).

## Disclosures

None of the authors have potential conflicts of interest to be disclosed

## Notes

### Competing Interest Statement

The authors have declared no competing interest.

### Clinical Trial

The data were collected from the archived database, therefore this study is not registered as a clinical trial.

### Author Declarations

This study is approved by Dutch committee on research involving human participants on 28th March 2017 [De Medisch Ethische Toetsingscommissie van Kempenhaeghe (17.04)].

### Summary of Updates

Abbreviation description in Table 1 added

