## Appendices for "Pitfalls in EEG analysis in patients with non-convulsive status epilepticus"

**Appendix A.** Annotation agreement code.

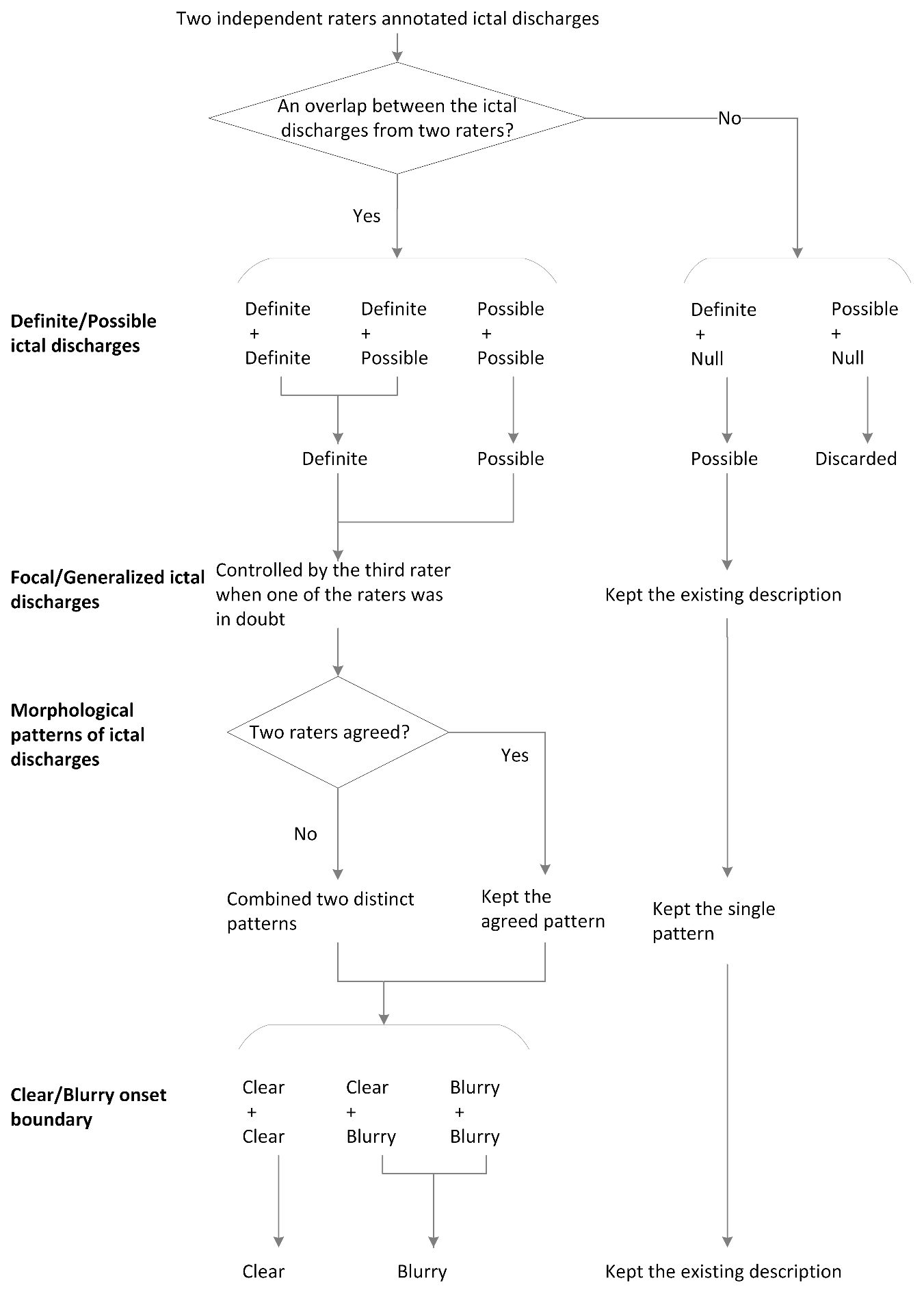

**Appendix B.** Steps of the self-developed system for automated EEG analysis in the monitoring of patients with NCSE.

| **Steps for analyzing EEG signals in our self-developed system** | | **Remarks** |
| --- | --- | --- |
| 1. Preprocessing | 1.1 Band-pass filtered [0.5-45 Hz]; 1.2 Subtract the average of all channel data from each channel data | To reduce interference caused by muscle or movement artefacts and direct current offsets. |
| 2. Feature extraction | 2.1 Commonly used time-frequency features in ictal detection: (Wang *et al.*, 2017; Aldana *et al.*, 2019) - approximation and detail coefficients corresponding to brain wave bands via wavelet decomposition analysis using wavelet Daubechies 4; - power spectrum within each brain wave band; - sample entropy; - distances between signal peaks and troughs | Brain wave bands we used:  delta wave:  0.5-3 Hz;  theta wave:  3-8 Hz;  alpha wave:  8-13 Hz;  beta wave:  13-30 Hz;  gamma wave:  30-45 Hz |
| 3. Classification | 3.1 Classified the EEG epochs into ictal discharge and normal activity using a binary RUSBoost classifier:   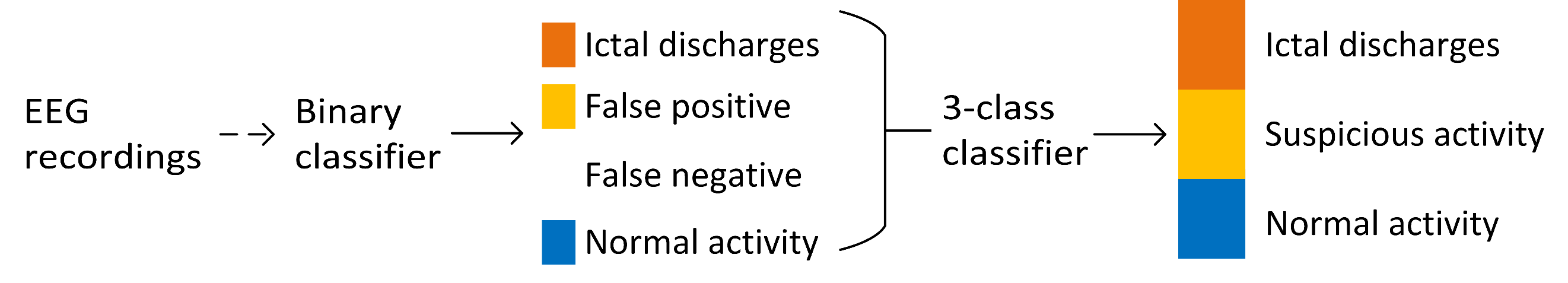   3.2 Created a synthetic 3-class RUSBoost classifier from the results of the binary classifier; 3.3 Used the synthetic 3-class classifier to classify EEG epochs into three categories: ictal discharges, suspicion activity, and normal activity | RUSBoost classifier extends AdaBoost classifier in addressing class-imbalance problems(Seiffert *et al.*, 2010) |

EEG: electroencephalography; NCSE: non-convulsive status epilepticus.

Aldana YR, Hunyadi B, Reyes EJM, Rodriguez VR and Van Huffel S. Nonconvulsive epileptic seizure detection in scalp eeg using multiway data analysis. IEEE J Biomed Heal Informatics 2019; 23(2): 660–671. doi: 10.1109/JBHI.2018.2829877.

Seiffert C, Khoshgoftaar TM, Van Hulse J and Napolitano A. RUSBoost: a hybrid approach to alleviating class imbalance. IEEE Trans Syst Man, Cybern - Part A Syst Humans 2010; 40(1): 185–197. doi: 10.1109/TSMCA.2009.2029559.

Wang L, Arends JBAM, Long X, Cluitmans PJM and van Dijk JP. Seizure pattern-specific epileptic epoch detection in patients with intellectual disability. Biomed Signal Process Control 2017; 35: 38–49. doi: 10.1016/j.bspc.2017.02.008.

**Appendix C.** The detailed characteristics of the individual subjects.

**Table 1.** NCSE patient demography

| **Patients** | **Intellectual disability:**  **normal**  **above 70;**  **light**  **(50-70); moderate (30–50); severe**  **(0–30)** | **Recording duration** | **Recorded sleep period** | **Preexisting epileptic encephalopathy** | **Clinical signs during recordings and seizure histories** | **Remarks** |
| --- | --- | --- | --- | --- | --- | --- |
| 1 | Light | Continuous  46 minutes | Obscured by epileptic activity (cannot be seen anymore) | Yes | Some myoclonias and showed panic with impaired awareness | Not applicable |
| 2 | Severe | Continuous  39 minutes | Not recorded | No | Tonic-clonic seizure with impaired awareness; atypical absence seizure with impaired awareness; myoclonic or atonic seizures with retained awareness | Not applicable |
| 3 | Severe | Continuous  46 minutes | Not recorded | Yes | Absences with impaired awareness; atonic seizures with impaired awareness;  tonic seizures with impaired awareness | Kabuki Syndrome |
| 4 | Severe | Continuous  8 hours and 50 minutes | Recognizable without specific sleep phenomena (not obvious sleep stages) | No | Laughing with impaired awareness; Tonic-clonic seizure with impaired awareness; NCSE with impaired awareness | Cardio-facio-cutaneous syndrome |
| 5 | Light | Continuous  2 hours and 53 minutes | Not recorded | No | Myoclonic and atonic seizures and loss of urine with impaired awareness | Not applicable |
| 6 | Severe | Continuous  22 hours and 19 minutes | Normal sleep but many seizures during sleep | No | Tonic-clonic seizures or tonic seizures with impaired awareness | Not applicable |
| 7 | Moderate | Continuous  21 hours and 45 minutes | Normal sleep | No | Myoclonias and automatisms seizures with impaired seizures; myoclonic and clonic seizures with impaired awareness; tonic seizures with impaired awareness | Not applicable |
| 8 | Normal | Non-continuous  50 hours and 50 minutes | Normal sleep | No | Focal seizure evolving into NCSE with impaired awareness | Ring chromosome 20 syndrome |
| 9 | Normal | Non- continuous 24hours | Normal sleep | No | Focal seizure evolving into tonic-clonic seizures with impaired awareness; two of six recorded seizures evolving into NCSE | Not applicable |
| 10 | Light | Non- continuous 25 hours and 24 minutes | 10% to 50% abnormal sleep due to the high frequency inter-ictal epileptic spikes and also normal sleep without spikes | No | Myoclonias with automatic behavior with impaired awareness; tonic-clonic seizures with impaired awareness; atypical absence (1-minute long) with impaired awareness; aphatic seizure with speak ability and retained awareness | Developmental encephalopathy and Genetic encephalopathy |
| 11 | Light | Non- continuous 25 hours and 56 minutes | 50% to 85% abnormal sleep due to the high frequency inter-ictal epileptic spikes and also normal sleep without spikes | No | NCSE with impaired awareness; myoclonic seizures with impaired awareness; tonic seizures with impaired awareness | Not applicable |
| 12 | Normal | Non- continuous 21 hours and 5 minutes | Normal sleep | No | Myoclonus, automatic behavior and vocalization with impaired awareness; NCSE evolved into tonic-clonic seizures with impaired awareness | Not applicable |
| 13 | Normal | Non- continuous 23 hours and 36 minutes | Not recorded | No | NCSE evolved into tonic-clonic seizures with impaired awareness; eyelid myoclonias preceding a tonic-clonic seizure;  Tonic-clonic seizure due to withdrawal anti-epileptic drugs | Not applicable |
| 14 | Normal | Non- continuous 24 hours and 25 minutes | Normal sleep | No | Tonic-clonic seizure with impaired awareness; atypical absences evolved into NCSE (1 hour and 15 minutes) with impaired awareness | Not applicable |
| 15 | Normal | Non- continuous 23 hours and 55 minutes | Normal sleep | No | Tonic-clonic seizures evolved into NCSE with impaired awareness | Not applicable |
| 16 | Normal | Non- continuous 23 hours and 23 minutes | Normal sleep | No | NCSE with slow reactions with impaired awareness; myoclonic-atonic seizures with retained awareness | Not applicable |

NCSE: non-convulsive status epilepticus.

**Table 2.** Suspected NCSE patient demography

| **Patients** | **Intellectual disability:**  **normal**  **above 70;**  **light**  **(50-70); moderate (30–50); severe**  **(0–30)** | **Recording duration** | **Recorded sleep period** | **Preexisting epileptic encephalopathy** | **Clinical signs during recordings and seizure histories** | **Remarks** |
| --- | --- | --- | --- | --- | --- | --- |
| 1 | Light | Continuous ca. 13 minutes | Not recorded | No | Myoclonic absence with impaired awareness | Not applicable |
| 2 | Moderate | Continuous ca. 18 minutes | Not recorded | Yes | No | FIRDA |
| 3 | Severe | Continuous  ca. 28 minutes | Unknown: difficult to distinguish between sleep and awake | Yes | Unknown (no seizure) | Not applicable |
| 4 | Normal | Continuous ca. 21 minutes | Not recorded | Yes | Unknown (no seizure) | Not applicable |
| 5 | Moderate | Continuous ca. 50 minutes | Sleep was recorded with many epileptiform (>85%) activities without status | Yes | Unknown (no seizure) | Not applicable |
| 6 | Severe | Continuous ca. 1 hour 2 minutes | Short normal sleep | No | Tonic seizures with impaired awareness | Severe mentally impaired with normal EEG |
| 7 | Normal | Continuous ca. 16 minutes | Not recorded | No | Focal seizures from left temporal lobe with impaired awareness | Not applicable |
| 8 | Normal | Continuous ca. 11 minutes | Not recorded | No | Unknown (no seizure) | Not applicable |
| 9 | Light | Continuous ca. 21 minutes | Normal sleep and slow wave sleep | Yes | Unknown (no seizure) | Not applicable |
| 10 | Severe | Continuous ca. 16 minutes | Not recorded | No | Two short absences with impaired awareness | Not applicable |
| 11 | Severe | Continuous ca. 5 minutes | Non-REM sleep patterns | Yes | Myoclonic seizure with retained awareness and motor signs; tonic seizure with retained awareness; absences with impaired awareness | High proportion of inter-ictal epileptic activities. |
| 12 | Normal | Continuous ca. 33 minutes | Short normal sleep | No | Status biPLD with impaired awareness and no-motor signs | Not applicable |

biPLD: bilateral period laterized discharges; EEG: electroencephalography; FIRDA: frontal intermittent rythmicdelta activity ; NCSE: non-convulsive status epilepticus; REM: rapid eye movement.

**Appendix D.** The interrater agreement of the individual subjects.

| Subject | Cohen’s kappa | Fleiss’s kappa |
| --- | --- | --- |
| 1 | 1.00 | 1.00 |
| 2 | -0.14 | 0 |
| 3 | 0 | 0 |
| 4 | 0.32 | 0.47 |
| 5 | 0.64 | 0.65 |
| 6 | 0.69 | 0.89 |
| 7 | 0.25 | 0.51 |
| 8 | 0.21 | 0.49 |
| 9 | 0.60 | 0.66 |
| 10 | 0 | 0.01 |
| 11 | 0.33 | 0.51 |
| 12 | 0.18 | 0.28 |
| 13 | 0.01 | 0.01 |
| 14 | 0.48 | 0.48 |
| 15 | 0.73 | 0.74 |
| 16 | -0.02 | 0 |

| **Subject** | **Remarks on the individual subjects about EEG over-interpretation** |
| --- | --- |
| 1 | Only one status epileptic; no extra remarks from the neurologists |
| 4 | Many potential false positives due to varied slow waves, and it was difficult for human raters to annotate ictal discharges |
| 5 | Focal slow waves were the hallmark of the seizures, intermixed with some spike waves; some slow activities during drowsiness disturbed the accuracy of ictal annotation |
| 6 | No extra remarks from the neurologists |
| 7 | No extra remarks from the neurologists |
| 8 | Same as the conclusion of subject 5 |
| 9 | The algorithm detected both ictal and abnormal activities |
| 10 | Too many inter-ictal discharges, human raters had difficulties to distinguish ictal discharges from this recording |
| 11 | There were several spike-wave concentrations with a high proportion of slow activities and inter-ictal activities, it was doubtful to determine ictal or inter-ictal activities |
| 12 | Frequency of sharp waves in the ictal periods was a bit higher than the inter-ictal periods, which may lead to false positive events by the algorithm |
| 13 | A tonic-clonic seizure and some abnormal activities were observed in this this recording |
| 14 | Many short ictal discharges in this EEG recording, the annotated periods look not very typical “ictal” |
| 15 | High agreement between annotations and the outputs of the algorithm; no extra remarks from the neurologists |
| 16 | Abnormal EEG activities and no definite ictal discharges. This recording is not suitable for the algorithm, given its extremely low inter-agreement |

**Appendix E.** The individual subject remarks about EEG over-interpretation and their proportions.

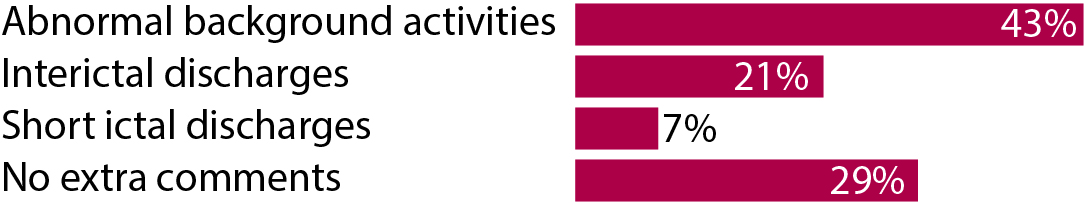

EEG: electroencephalography.
